# Exercise Capacity and Mental Health in Adults With a Systemic Right Ventricle

**DOI:** 10.64898/2026.08.24.26361239

**Authors:** Bryan P Mosher, Jennifer P Woo, Jeffrey W Christle, Jason V Tso, Euan A Ashley, Daniel E Clark

**Author notes:** Address for correspondence: Bryan P Mosher, Division of Cardiovascular Medicine, Adult Congenital Heart Disease Program, Stanford University, 300 Pasteur Drive, 3rd Floor, Room A32, Stanford, CA 94305, USA. Social media: X: @bryanmosher09; Instagram: bryanm098516; LinkedIn public profile URL: www.linkedin.com/in/bryan-mosher-md-phd-577107336. Adults with a systemic right ventricle (sRV) face interconnected physiologic and psychosocial challenges. d-TGA after atrial switch was associated with lower exercise capacity, reduced sRV function, and greater mental health burden than ccTGA.

## Abstract

**Background:** Adults with a systemic right ventricle (sRV) due to congenitally corrected transposition of the great arteries (ccTGA) or atrial switch repair for d-transposition of the great arteries (d-TGA) experience substantial physiologic and psychosocial morbidity. Relationships among exercise capacity, sRV function, and mental health remain incompletely characterized.

**Objectives:** To characterize relationships among anatomic subtype, exercise capacity, sRV function, and mental health in adults with sRV physiology.

**Methods:** We performed a retrospective cohort study of adults with ccTGA or d-TGA (Mustard/Senning) followed at a tertiary Adult Congenital Heart Disease program from 2000 to 2025. Clinical, imaging, cardiopulmonary exercise testing, and patient-reported data were obtained from electronic health records. Mental health diagnoses were identified from clinical documentation. Functional status was assessed using NYHA class and the Kansas City Cardiomyopathy Questionnaire (KCCQ-12).

**Results:** Among 137 adults (ccTGA, n = 51; d-TGA, n = 86), percent-predicted peak VO2 was lower in d-TGA than ccTGA (60% vs 74%, p < 0.001), as was sRV systolic function (41 ± 11% vs 47 ± 10%, p < 0.01). Anxiety or depression was more common in d-TGA (46% vs 25%, p < 0.05). Across the cohort, anxiety or depression was associated with lower exercise capacity, worse NYHA functional class, and lower KCCQ scores.

**Conclusions:** Adults with d-TGA following atrial switch have lower exercise capacity, reduced sRV systolic function, and greater mental health burden than adults with ccTGA. These findings support integrated assessment of physiologic performance, functional status, and mental health in adults with sRV physiology.

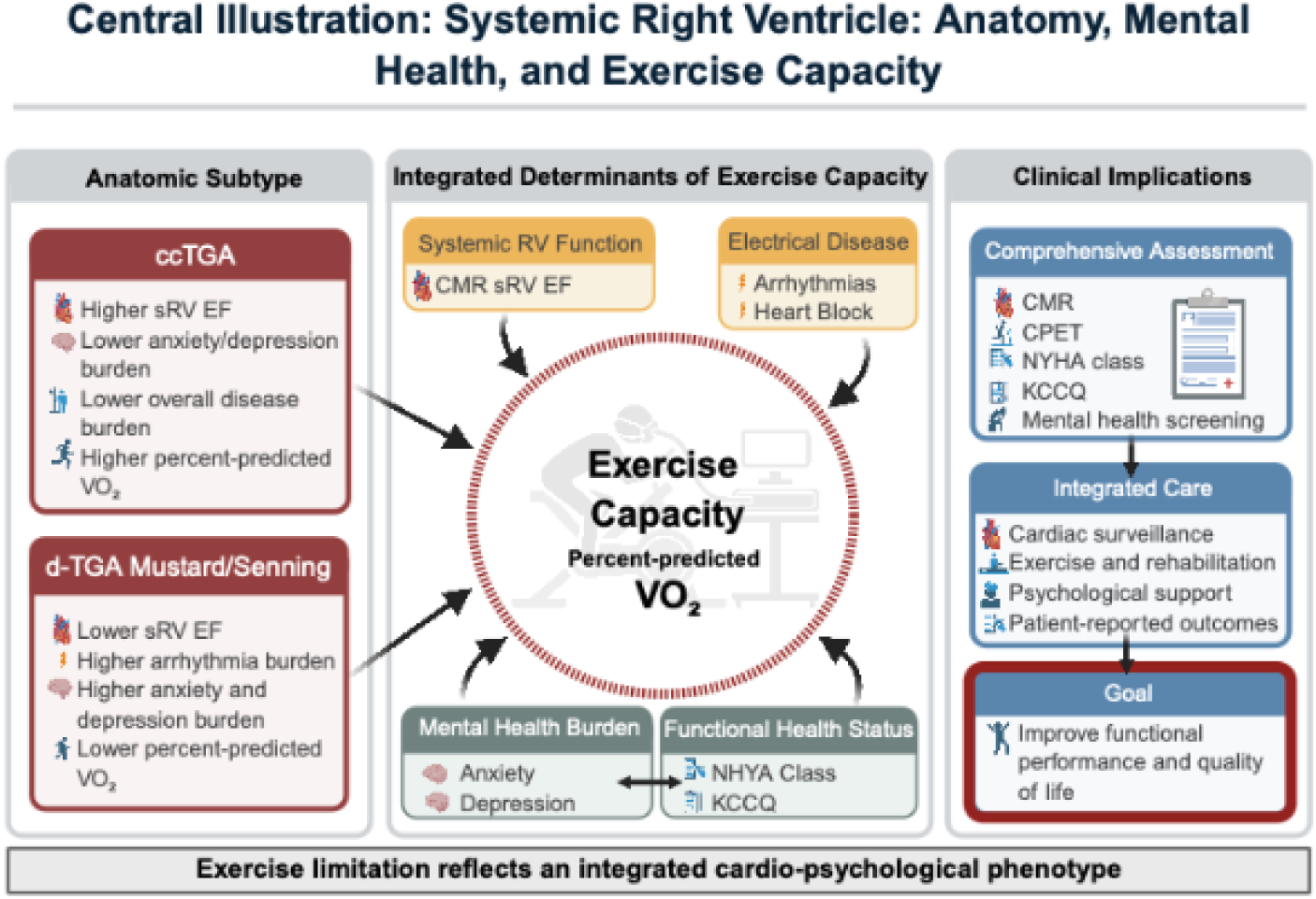

Central Illustration. Integrated Cardio-Psychological Determinants of Exercise Capacity in Adults With a Systemic Right Ventricle. Adults with a sRV exhibit distinct clinical profiles according to anatomic subtype. Patients with d-TGA following atrial switch repair generally demonstrate lower exercise capacity, reduced systemic right ventricular systolic function, and a greater burden of anxiety, depression, and arrhythmias than patients with ccTGA. Exercise capacity reflects an integrated phenotype influenced by ventricular function, electrical disease, mental health, and functional health status. These findings support comprehensive assessment incorporating cardiac imaging, cardiopulmonary exercise testing, patient-reported health status, and mental health screening to guide multidisciplinary management and optimize functional performance and quality of life. Abbreviations: ccTGA = congenitally corrected transposition of the great arteries; CMR = cardiac magnetic resonance; CPET = cardiopulmonary exercise testing; d-TGA = d-transposition of the great arteries; KCCQ = Kansas City Cardiomyopathy Questionnaire; NYHA = New York Heart Association; sRV = systemic right ventricle; VO2 = peak oxygen consumption.

## Introduction

Heart failure is a growing cause of morbidity and mortality among adults with congenital heart disease (CHD), particularly among those with systemic right ventricular (sRV) physiology. Despite advances in surgical and medical care, adults with congenitally corrected transposition of the great arteries (ccTGA) and d-transposition of the great arteries (d-TGA) following atrial switch repair remain at increased risk for ventricular dysfunction, arrhythmias, heart failure, and premature mortality.^1–6^ Reduced exercise capacity is a hallmark manifestation of sRV disease and is strongly associated with adverse clinical outcomes and quality of life.^1,2,7^

Mental health disorders are increasingly recognized as important determinants of outcomes in adults with CHD.^8,9^ Anxiety and depression are associated with impaired quality of life,^10^ increased symptom burden, worse functional status, and adverse cardiovascular outcomes^11^ across several CHD populations.^12–15^ However, little is known about the relationship between mental health burden and exercise capacity in adults with sRV physiology, or whether these associations differ between patients with ccTGA and those with d-TGA following atrial switch repair. Defining the relationship between mental health burden and objective measures of functional capacity may improve risk stratification and identify opportunities for integrated interventions that address both physiologic impairment and psychological well-being in adults with sRV physiology.

Accordingly, we sought to characterize the relationships among anatomic subtype, exercise capacity, sRV systolic function, and mental health status in adults with sRV physiology. We hypothesized that anxiety and depression would be associated with lower exercise capacity, worse functional status, and poorer patient-reported health status, and that these associations would be most pronounced among adults with d-TGA following atrial switch repair.

## Methods

### Study design and population

A single-center retrospective cohort study was performed on 137 adults (≥18 years) from two anatomic groups: (1) ccTGA, defined by atrioventricular and ventriculo-arterial discordance confirmed by imaging, and (2) d-TGA after Mustard or Senning atrial switch surgery confirmed by surgical reports and imaging. Eligible patients were identified from January 2000 to December 2025 at Stanford University. Manual chart review was performed to confirm CHD diagnosis, demographic, and clinical data. Patients were excluded if they had undergone arterial switch repair, Rastelli procedure, double switch operation, or Fontan palliation. Individuals with incomplete exercise or imaging data were retained if sufficient clinical and mental-health data were available for comparison. The institutional review board approved this study (IRB 78353).

### Demographic information and clinical variables

Data were extracted from the Stanford electronic medical record (EMR). Demographic information included age, sex, and body mass index (BMI). Sex was obtained from the EMR as documented at the time of clinical care. Clinical history included congenital heart defect, cardiac surgical intervention (none, Mustard, Senning for the d-TGA population, and pulmonary artery band and valve repair for the ccTGA population), hypertension (HTN), chronic kidney disease (CKD), hypothyroidism, hemoglobin A1c, and device therapy (pacemaker or implantable cardioverter-defibrillator). Surgical data were confirmed from operative reports or prior cardiology summaries.

### Cardiac imaging

sRV function was quantified by transthoracic echocardiogram (TTE) and cardiac magnetic resonance (CMR) imaging when available. When performed at Stanford, TTE studies were done in accordance with standard congenital TTE protocols.^16^ CMR studies done at Stanford were performed on either a 1.5-Tesla (T) or 3T system (GE Healthcare, Milwaukee, Wisconsin; Optima 450 W and MR750) in accordance with standard CMR protocols for patients with CHD.^17^ When multiple TTE and CMR studies were available, the most recent scans were used.

### Cardiopulmonary exercise testing

Exercise capacity was assessed using symptom-limited CPET with metabolic gas analysis (COSMED Quark, COSMED Omnia) on a cycle ergometer (Ergoline). A ramp protocol was chosen to achieve 8-12 minutes of exercise duration. Continuous 12-lead ECG and blood pressure monitoring were performed. Percent-predicted peak VO2 (ppVO2) was calculated using standard equations adjusted for age, sex, and body size. Submaximal tests determined by respiratory exchange ratio (RER < 1.05) were excluded. When multiple CPETs were available, ppVO2 values were averaged to generate a patient-level mean for analysis.

### Functional class and health status

NYHA class was assigned by treating physicians at the time of most recent cardiology clinical visit. Patient-reported outcomes were previously assessed using the Kansas City Cardiomyopathy Questionnaire-12 (KCCQ). When multiple KCCQ tests were done, the most recent test was used. Domain scores (Physical Limitation, Symptom Frequency, Quality of Life, Social Limitation) and Overall Summary Score (OSS) were recorded; higher values indicate better perceived health.

### Mental health assessment

Anxiety and depression were identified based on (1) diagnosis in clinical notes, or (2) prescription for anxiolytic or antidepressant medication. Medication alone without documentation was not counted unless clearly prescribed for psychiatric indication.

### Statistical analysis

Continuous variables are presented as median [interquartile range] or mean ± standard deviation based on distribution assessed by the Shapiro-Wilk test. Between-group comparisons were made using Wilcoxon rank-sum or Student’s t-tests as appropriate. Categorical variables were compared using chi-square or Fisher’s exact tests. Spearman correlation coefficients were used to evaluate associations among physiologic and psychosocial variables (ppVO2, sRV EF, NYHA class, KCCQ score), given their non-normal or ordinal distributions.

Multivariable linear regression models were constructed to evaluate the association between ccTGA and d-TGA after Mustard/Senning and (1) ppVO2 and (2) KCCQ overall summary score. Covariates selected a priori included anatomic subtype, age, sex, sRV EF on CMR, and diagnosis of anxiety or depression. Analyses were performed using R version 2024.12.1+563, and results are reported as β coefficients with 95% confidence intervals, and statistical significance was defined as p < 0.05.

## Results

### Cohort characteristics

A total of 137 adults with sRV physiology met inclusion criteria, including 51 patients with ccTGA and 86 patients with d-TGA following Mustard or Senning repair. In the combined cohort, median age at most recent follow-up was 36 [29–43] years, and 84 patients (61%) were male. When stratified by anatomy, age and sex distribution did not differ significantly between ccTGA and d-TGA groups. Compared with ccTGA, patients with d-TGA demonstrated a greater burden of cardiovascular comorbidities, including arrhythmia, hypertension, chronic kidney disease, and abnormal glycemic status. Baseline demographic and clinical characteristics are summarized in Table 1.

**Table 1.**
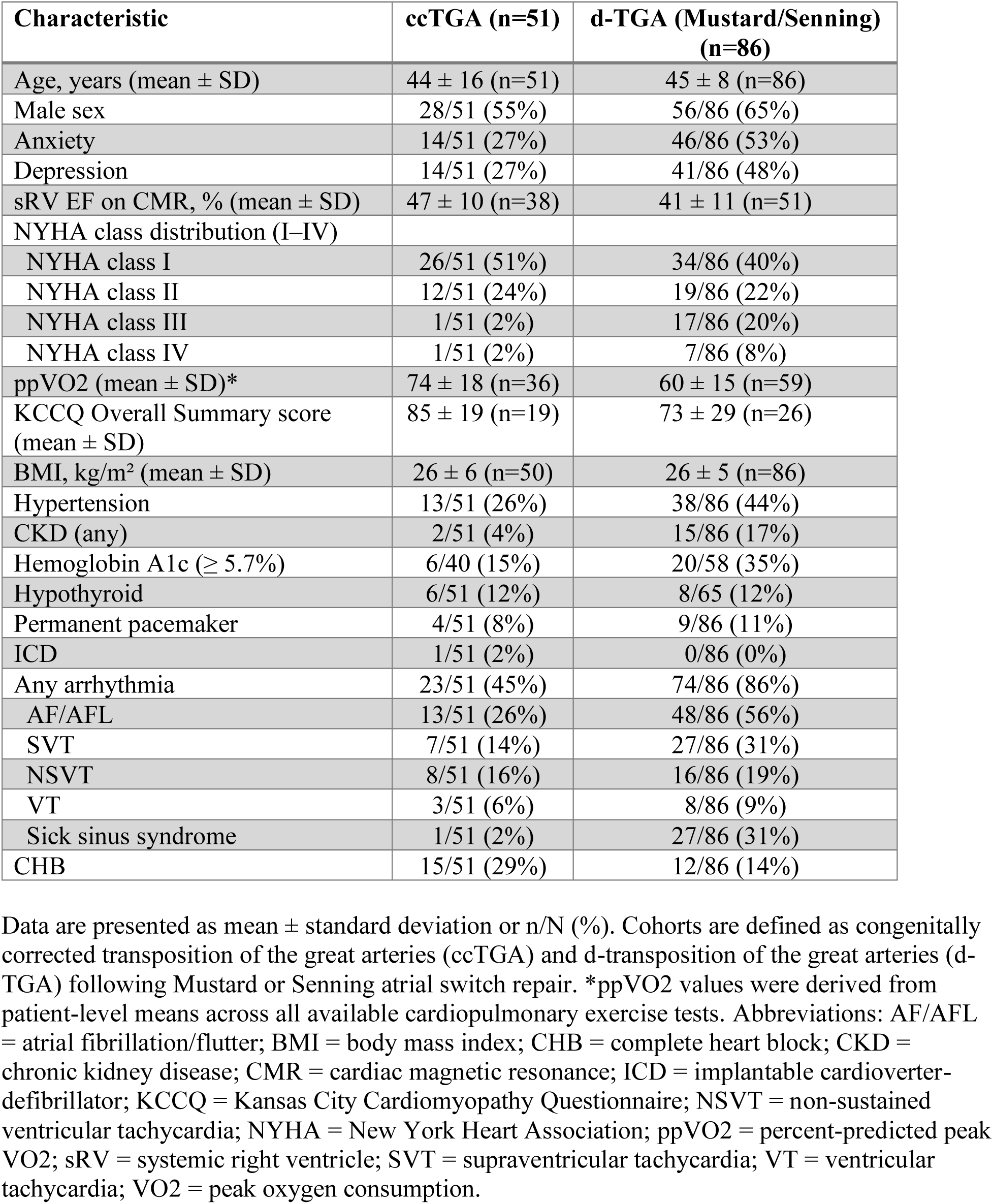
Baseline Characteristics.

### Exercise capacity and mental health

CPET data were available for 95 patients in the combined cohort. Across all sRV patients, ppVO2 was reduced, with a mean of 66 ± 18%. When stratified by anatomy, d-TGA patients demonstrated lower ppVO2 compared with ccTGA patients (60 ± 15% vs 74 ± 18%, p < 0.001; Figure 1). In the combined sRV cohort, anxiety was documented in 60 patients (44%) and depression in 55 patients (40%). Both anxiety and depression were more prevalent among d-TGA patients compared with ccTGA patients (anxiety: 53% vs 27%, p < 0.01; depression: 48% vs 27%, p < 0.05). Patients with documented anxiety or depression demonstrated lower ppVO2 compared with patients without either diagnosis (p < 0.05, Figure 2). When stratified by anatomy, this association was observed among d-TGA patients, in whom those with anxiety or depression had lower ppVO2 than those without these diagnoses (p < 0.05). In contrast, no significant differences in ppVO2 by mental health status were observed among ccTGA patients (Figure 3). BMI did not differ according to anxiety status (median 24.8 [IQR 21.8-27.5] vs 25.7 [22.3-29.9] kg/m², p = 0.166) or depression status (median 24.6 [21.8-27.5] vs 25.7 [22.4-30.3] kg/m², p = 0.105). Furthermore, BMI was not associated with mean ppVO2 (Spearman ρ = -0.01, p = 0.947).

**Figure 1.**
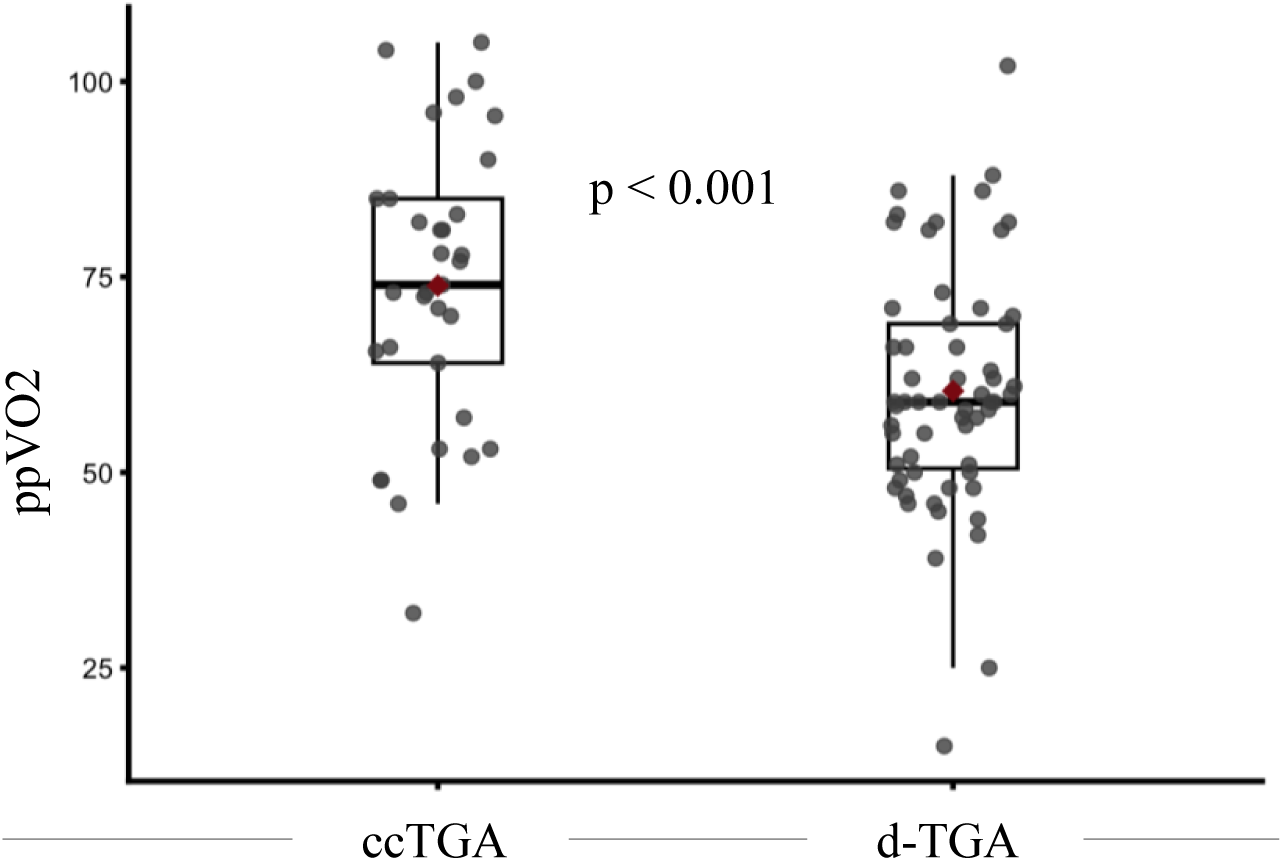
Exercise Capacity by Anatomic Subtype. ppVO2 is shown for adults with a sRV, stratified by anatomic subtype. Patients with d-TGA following atrial switch (Mustard/Senning; n = 59) demonstrated lower exercise capacity compared with those with ccTGA (n = 36; 60 ± 15% vs 74 ± 18%, p < 0.001). Each point represents an individual patient, with values calculated as the mean of all available cardiopulmonary exercise tests per patient. Box plots display the median and interquartile range, with whiskers representing 1.5 times the interquartile range. Red markers indicate group means. Statistical comparison was performed using the Wilcoxon rank-sum test. Abbreviations: ccTGA = congenitally corrected transposition of the great arteries; d-TGA = d-transposition of the great arteries; ppVO2 = percent-predicted peak VO2; sRV = systemic right ventricle; VO2 = peak oxygen consumption.

**Figure 2.**
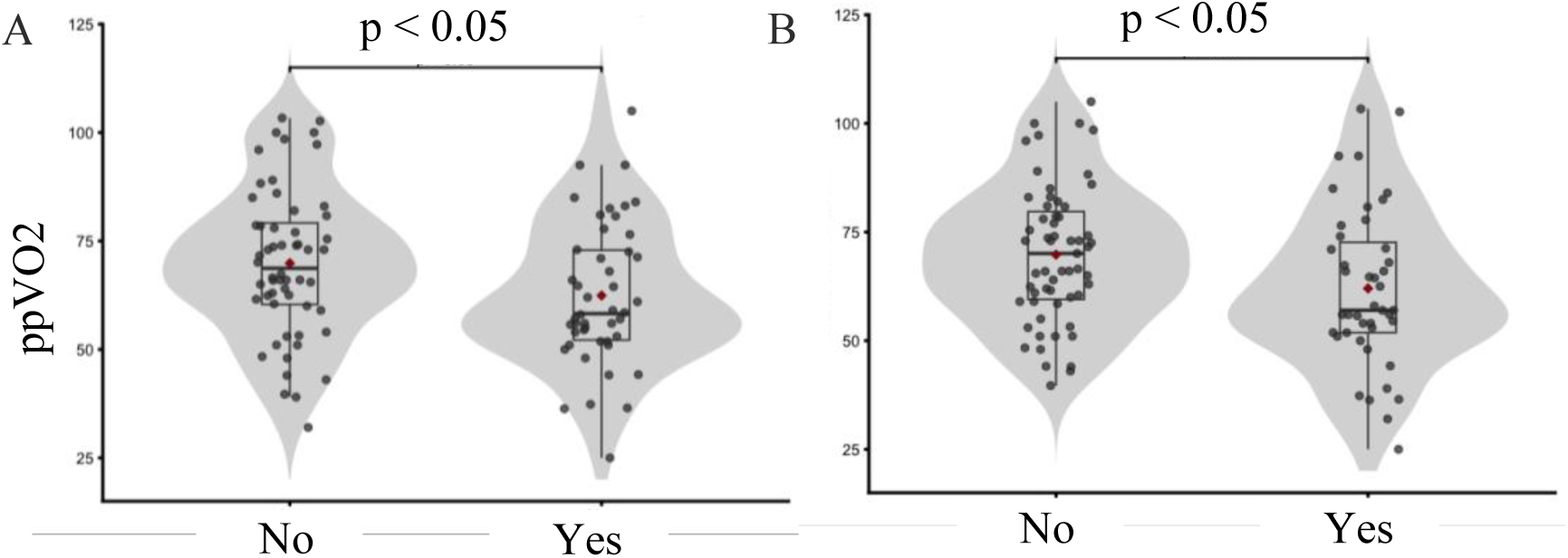
Association of Mental Health Status with Exercise Capacity. **(A)** ppVO2 in patients with and without a diagnosis of anxiety. **(B)** ppVO2 in patients with and without a diagnosis of depression. In the combined sRV cohort, patients with anxiety or depression demonstrated lower exercise capacity compared with those without these diagnoses (p < 0.05 for both comparisons). Each point represents an individual patient, with values calculated as the mean of all available cardiopulmonary exercise tests per patient. Violin plots illustrate the distribution of values, with overlaid box plots showing the median and interquartile range. Red markers indicate group means. Statistical comparisons were performed using the Wilcoxon rank-sum test. Abbreviations: ppVO2 = percent-predicted peak VO2; sRV = systemic right ventricle.

**Figure 3.**
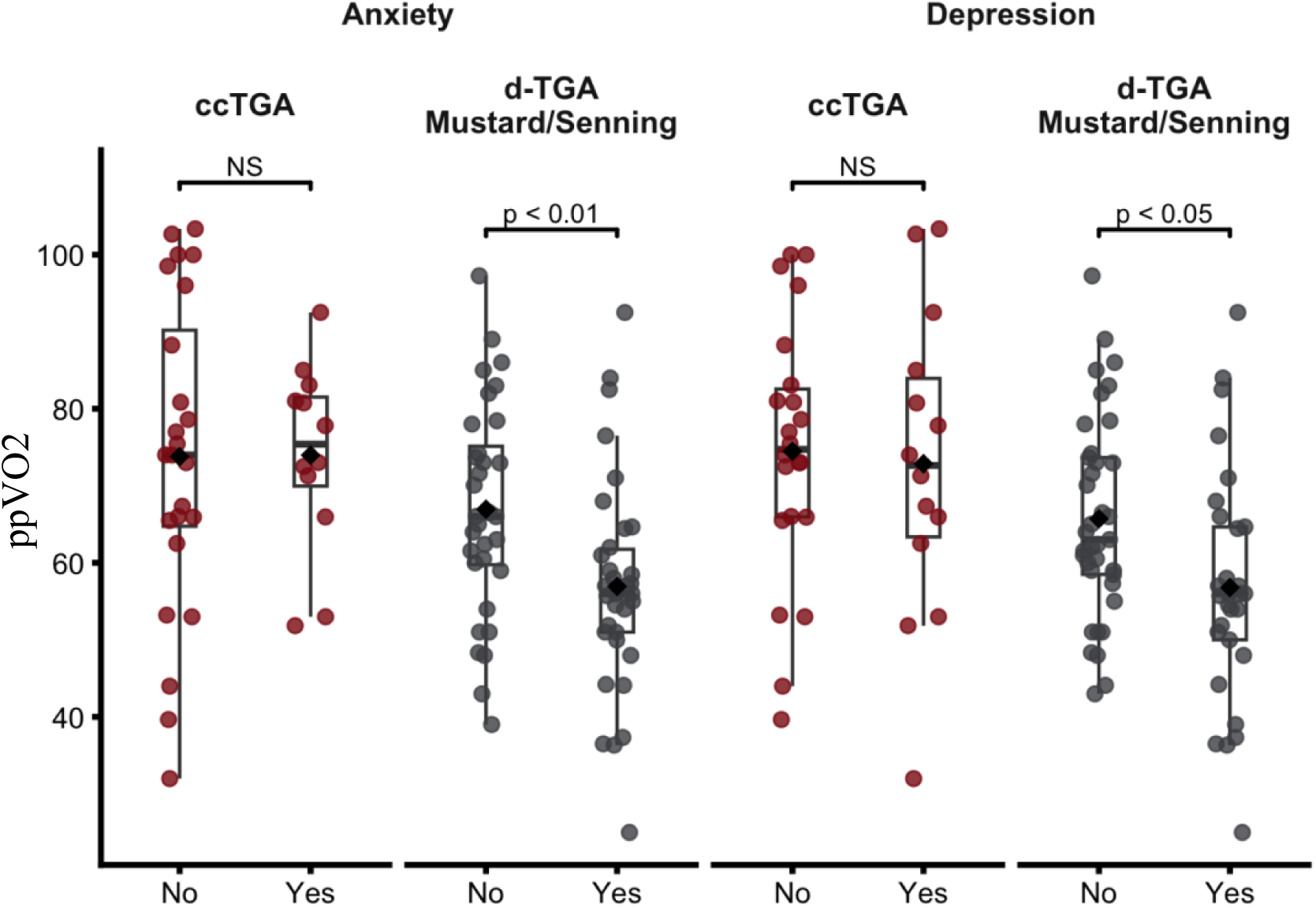
Exercise Capacity by Anxiety and Depression Status in ccTGA and d-TGA Patients. Box-and-scatter plots showing ppVO2 across groups stratified by anxiety and depression status. Data are presented for ccTGA and d-TGA following Mustard/Senning arial switch, with participants categorized as “No” or “Yes” for anxiety or depression. Individual data points are overlaid on box plots representing median and interquartile range. For both anxiety and depression panels, no significant differences were observed in ccTGA groups (NS). In contrast, participants with d-TGA Mustard/Senning who reported anxiety or depression showed significantly lower ppVO2 compared to those without these conditions (anxiety: p < 0.01; depression: p < 0.05). Abbreviations: ccTGA = congenitally corrected transposition of the great arteries; d-TGA = d-transposition of the great arteries; NS = not significant; ppVO2 = percent-predicted peak VO2.

### Functional class and patient-reported health status

Across the combined cohort, ppVO2 declined progressively with worsening NYHA functional class (Figure 4A). This pattern was present in both anatomic groups. Within individual NYHA classes, no statistically significant differences in ppVO2 were observed between ccTGA and d-TGA patients. In the combined cohort, patients with documented anxiety or depression were more likely to be classified in higher NYHA functional classes compared with patients without these diagnoses (Figure 4B). Higher KCCQ overall summary scores were associated with higher ppVO2 in both anatomic groups. Between-group comparisons demonstrated significantly higher ppVO2 in ccTGA compared with d-TGA patients within the highest KCCQ quartile, while no significant difference was observed between groups in the intermediate KCCQ range (Q2 50–74, Figure 4C).

**Figure 4.**
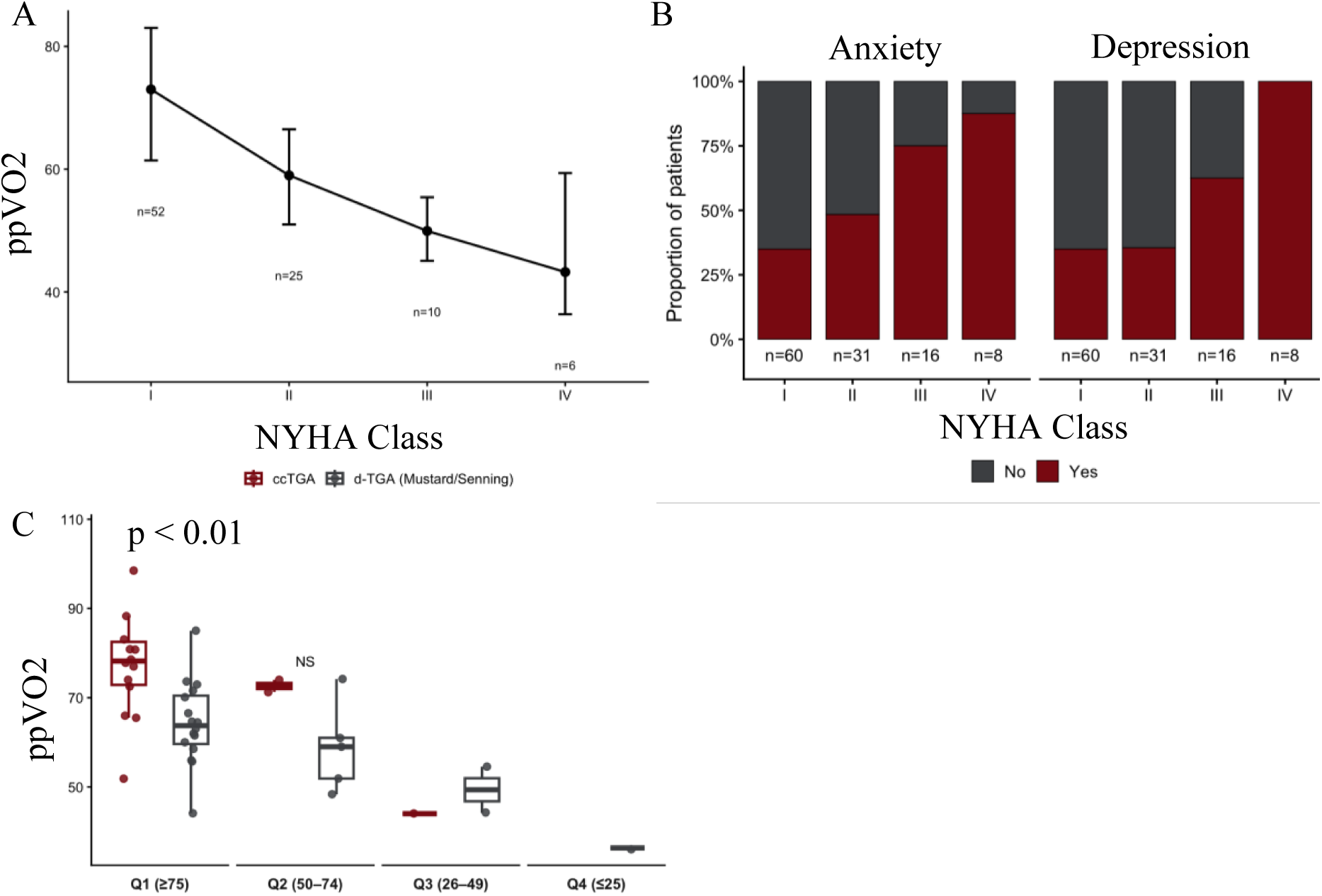
Functional Class, Mental Health, and Patient-Reported Health Status in Relation to Exercise Capacity. **(A)** ppVO2 by NYHA functional class in the combined cohort. ppVO2 declined progressively with worsening NYHA functional class. **(B)** Prevalence of anxiety and depression across NYHA functional classes in the combined cohort. Patients with anxiety or depression were more frequently represented in higher NYHA classes. **(C)** ppVO2 by KCCQ overall summary score quartiles, stratified by anatomic subtype. Higher KCCQ scores were associated with greater exercise capacity, with ccTGA patients demonstrating higher ppVO2 than d-TGA Mustard/Senning patients in the highest KCCQ quartile. ppVO2 values represent the mean of all available cardiopulmonary exercise tests per patient. In panel A, points and error bars represent group means ± standard deviation, with statistical comparison across NYHA classes performed using the Kruskal–Wallis test. In panel B, bars represent the proportion of patients with and without anxiety or depression within each NYHA class. In panel C, box plots display the median and interquartile range with whiskers representing 1.5 times the interquartile range; red elements denote ccTGA and gray elements denote d-TGA Mustard/Senning. Statistical comparisons between anatomic groups within KCCQ quartiles were performed using the Wilcoxon rank-sum test. Abbreviations: KCCQ = Kansas City Cardiomyopathy Questionnaire; NYHA = New York Heart Association; ppVO2 = percent-predicted peak VO2.

### Systemic right ventricular function

In the combined ccTGA and d-TGA cohort, patient-level mean ppVO2 differed across sRV EF categories by CMR (59 % in patients with EF ≤ 40 %, 75 % in those with EF 41–50 %, and 74 % in those with EF > 50 %, with a significant overall difference across EF categories on Kruskal–Wallis testing, p < 0.05; Figure 5A). When stratified by anatomy, ccTGA patients exhibited higher median sRV EF compared with d-TGA patients following atrial switch (p < 0.01; Figure 5B). CMR-derived sRV EF was positively correlated with ppVO2 in ccTGA patients (p < 0.05), whereas no significant correlation was observed in d-TGA patients palliated with Mustard or Senning procedures (Figure 5C). In the combined cohort, patients with anxiety demonstrated lower median sRV EF compared with those without anxiety (p < 0.05), whereas no difference was observed according to depression status (Figure 6A). However, when stratified by anatomy, sRV EF did not differ according to anxiety or depression status within either anatomic subgroup (Figure 6B). Between anatomic groups, sRV EF remained lower in d-TGA than in ccTGA irrespective of depression status (p < 0.05; Figure 6C).

**Figure 5.**
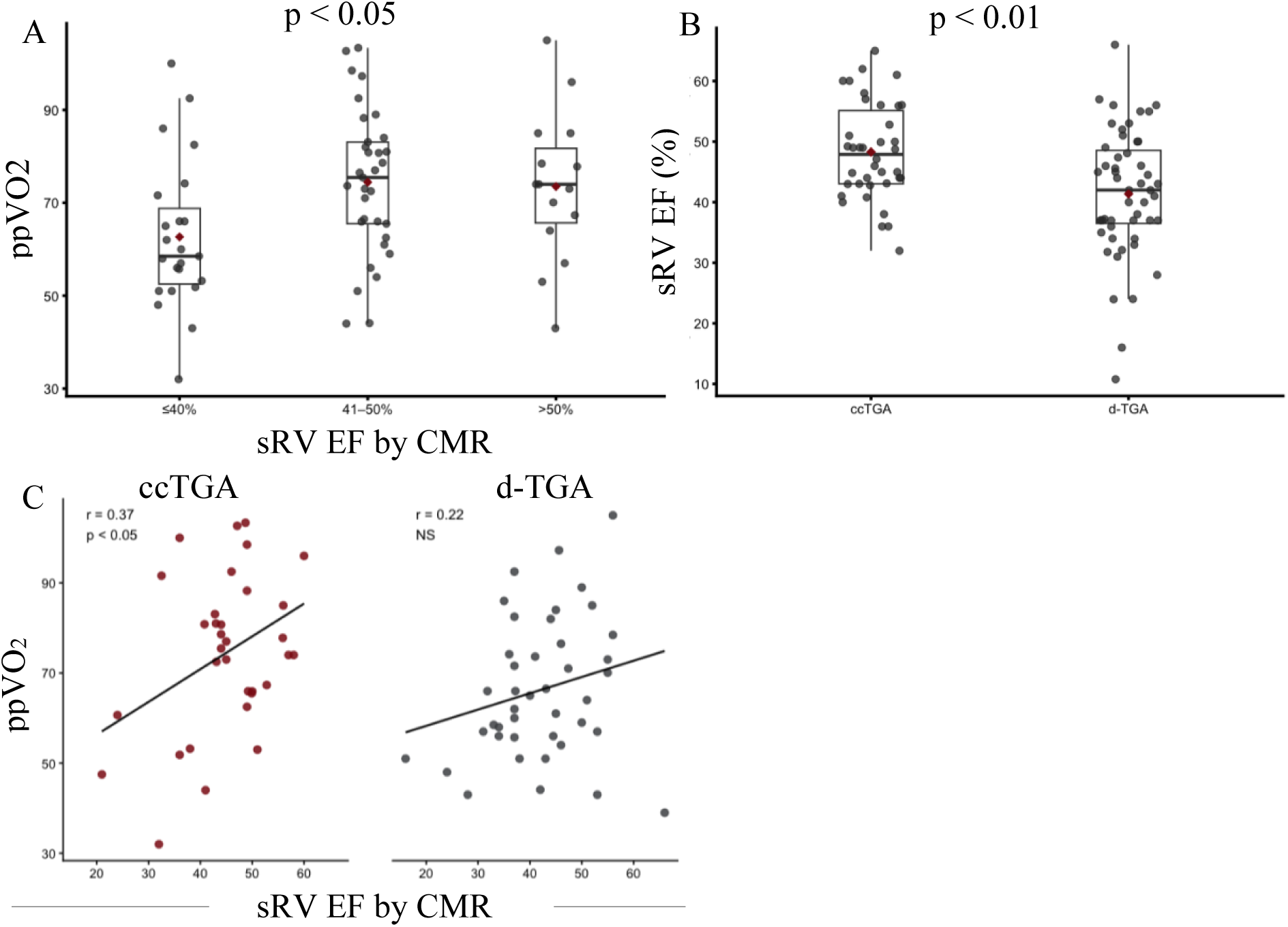
Systemic Right Ventricular Function and Exercise Capacity. **(A)** ppVO2 across categories of sRV EF measured by CMR in the combined cohort. **(B)** Distribution of CMR-derived sRV EF by anatomic subtype. **(C)** Relationship between ppVO2 and CMR-derived sRV EF stratified by anatomic subtype. ppVO2 differed across sRV EF categories, with higher values observed at greater levels of ventricular function. Patients with ccTGA demonstrated higher sRV EF compared with those with d-TGA following atrial switch. A positive association between sRV EF and ppVO2 was observed in ccTGA, whereas no significant relationship was present in d-TGA Mustard/Senning. ppVO2 values represent the mean of all available cardiopulmonary exercise tests per patient. In panel A, box plots display the median and interquartile range with whiskers representing 1.5 times the interquartile range; comparisons across EF categories were performed using the Kruskal–Wallis test. In panel B, box plots illustrate the distribution of CMR-derived sRV EF by anatomic subtype, with comparisons performed using the Wilcoxon rank-sum test. In panel C, each point represents an individual patient; lines indicate linear regression fits within each anatomic group. Statistical significance for correlations was assessed using Spearman rank correlation. Abbreviations: CMR = cardiac MRI; EF = ejection fraction; ppVO2 = percent-predicted peak VO2; sRV = systemic right ventricle.

**Figure 6.**
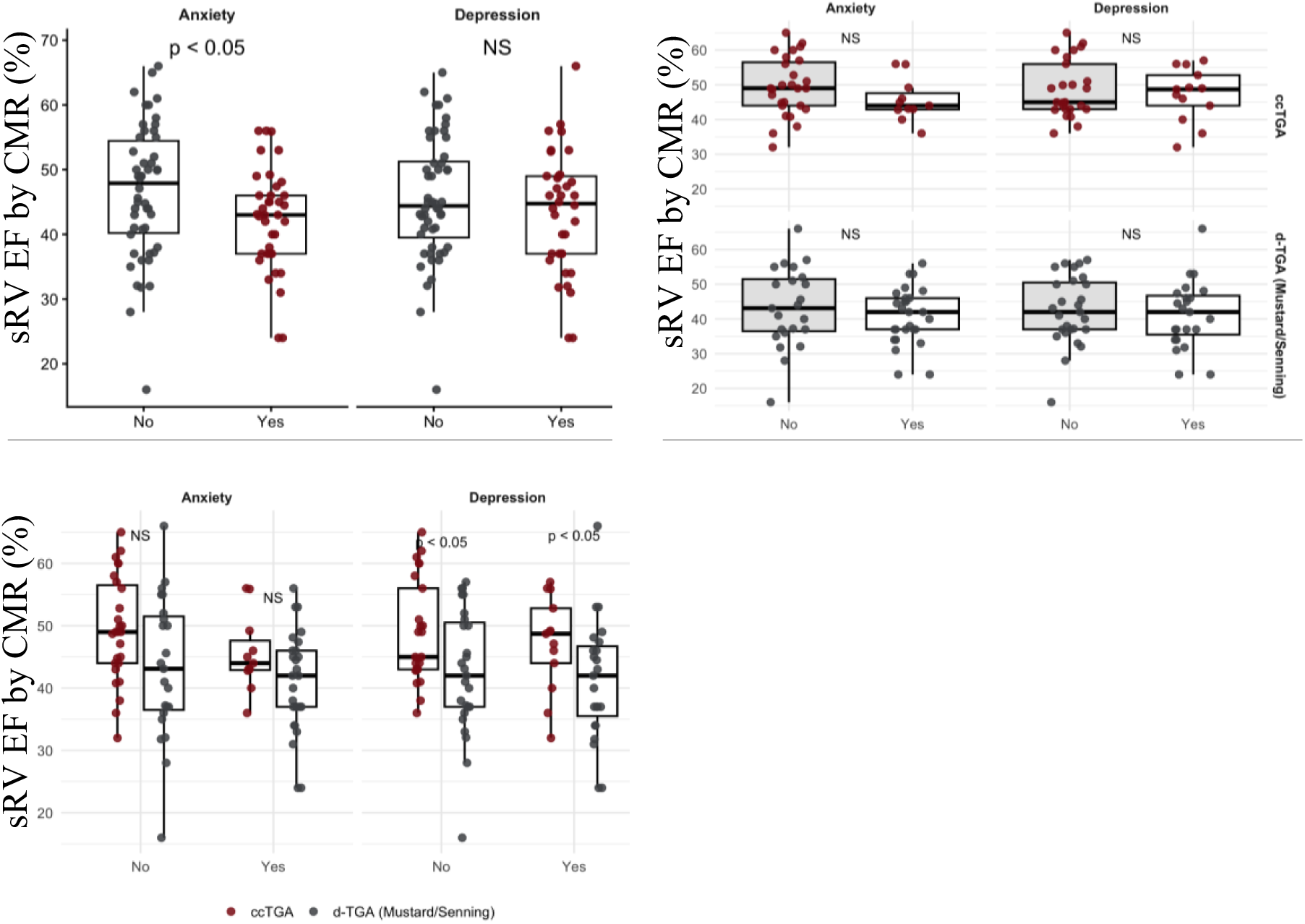
Mental Health Status and Systemic Right Ventricular Function. **(A)** CMR-derived sRV EF by anxiety and depression status in the combined cohort. **(B)** CMR-derived sRV EF by anxiety and depression status stratified by anatomic subtype. **(C)** CMR-derived sRV EF comparing anatomic subtypes within strata of anxiety and depression status. In the combined cohort, patients with anxiety demonstrated lower sRV EF compared with those without anxiety, whereas no difference was observed by depression status. When stratified by anatomy, sRV EF did not differ by anxiety or depression status within either ccTGA or d-TGA Mustard/Senning. However, when comparing between anatomic subtypes, sRV EF was lower in d-TGA compared with ccTGA among patients both with and without depression. sRV EF was measured by CMR. In all panels, box plots display the median and interquartile range with whiskers representing 1.5 times the interquartile range; individual points represent patients. Red elements denote ccTGA and gray elements denote d-TGA Mustard/Senning. Statistical comparisons were performed using the Wilcoxon rank-sum test. Abbreviations: CMR = cardiac MRI; EF = ejection fraction; sRV = systemic right ventricle.

### Multivariable predictors of exercise capacity and health status

In multivariable linear regression analysis, ccTGA anatomy was independently associated with higher ppVO2 compared with d-TGA following atrial switch (β 8.29, 95% CI 0.74 to 15.80, p < 0.05). Age, sex, sRV EF, and mental health diagnosis were not independently associated with ppVO2. In contrast, no variables were independently associated with KCCQ overall summary score, and the overall model was not statistically significant (Table 2). The KCCQ analysis was limited by a smaller sample size due to missing data.

**Table 2.** Multivariable predictors of exercise capacity and health status.

| Predictor | ppVO2 $\beta$ (95% CI) | p-value | KCCQ Score $\beta$ (95% CI) | p-value |
| --- | --- | --- | --- | --- |
| ccTGA vs d-TGA after atrial switch | 8.29 (0.74 to 15.80) | p < 0.05 | 5.98 (-10.40 to 22.40) | NS |
| Age (per year) | -0.08 (-0.43 to 0.26) | NS | -0.03 (-0.74 to 0.69) | NS |
| Male sex | -6.16 (-14.00 to 1.67) | NS | -2.84 (-20.60 to 14.90) | NS |
| sRV EF on MR (per 1%) | 0.21 (-0.25 to 0.67) | NS | -0.65 (-1.93 to 0.62) | NS |
| Mental health diagnosis | -5.70 (-13.00 to 1.62) | NS | -7.99 (-23.30 to 7.37) | NS |

Values are presented as $\beta$ coefficients with 95% CI. Regression models included anatomic subtype, age, sex, sRV EF measured by CMR, and the presence of anxiety or depression. ppVO2 values were derived from patient-level means across all available cardiopulmonary exercise tests. Mental health diagnosis was defined as the presence of either anxiety or depression.
Abbreviations: CI = confidence interval; CMR = cardiac magnetic resonance; EF = ejection fraction; KCCQ = Kansas City Cardiomyopathy Questionnaire; ppVO2 = percent-predicted peak VO2; sRV = systemic right ventricle; VO2 = peak oxygen consumption.

## Discussion

In this single-center cohort of adults with sRV physiology the principal findings are threefold. First, anxiety and depression were highly prevalent among adults with sRV physiology and were associated with lower exercise capacity, worse functional status, and poorer patient-reported health status. Second, these associations were most pronounced among adults with d-TGA following atrial switch, who also demonstrated lower exercise capacity and reduced sRV function compared with adults with ccTGA. Third, despite these relationships, anatomic subtype remained the primary independent correlate of exercise capacity after multivariable adjustment. Collectively, these findings support the concept of sRV disease as an integrated cardio-psychological phenotype in which anatomy, ventricular adaptation, and psychological well-being jointly shape patient experience.

In multivariable analysis, however, anatomic subtype emerged as the primary independent correlate of exercise capacity. After adjustment for age, sex, sRV EF, and mental health diagnosis, ccTGA remained associated with significantly higher ppVO2, whereas sRV function and mental health status were no longer independently associated. This finding suggests that while ventricular dysfunction and anxiety/depression track closely with exercise limitation, they may represent markers of overall disease severity rather than independent determinants of reduced exercise capacity.

Prior studies comparing exercise capacity in ccTGA and d-TGA after Mustard/Senning populations have reported heterogeneous findings, with some demonstrating similar ppVO2,^18^ others suggesting reduced capacity in ccTGA,^19^ and relatively limited contemporary data directly supporting worse performance in the d-TGA atrial switch population. Our findings provide additional clarity in this context, demonstrating consistently lower ppVO2 in d-TGA patients following atrial switch compared with those with ccTGA. Importantly, this difference persisted after adjustment for age, sex, sRV function, and mental health status, suggesting that anatomic subtype is independently associated with exercise capacity. Notably, our cohort was older than that reported by Kempny et al., raising the possibility that progressive age-related decline in the atrial switch population may account for some of the discrepancy between studies. These results support the concept that the physiologic consequences of atrial switch circulation may confer greater limitations in exercise performance than those observed in ccTGA.

More broadly, chronic sRV pressure loading results in progressive ventricular remodeling characterized by hypertrophy, fibrosis, chamber dilation, and increasing tricuspid regurgitation. These structural changes occur alongside electrical remodeling and increasing arrhythmia burden, ultimately reducing contractile reserve and limiting exercise capacity.

Several anatomic and surgical factors may contribute to the observed differences between ccTGA and d-TGA populations. In d-TGA, atrial switch repair introduces extensive atrial suture lines and baffle pathways that are associated with sinus node dysfunction, atrial arrhythmias, and venous pathway abnormalities, all of which can limit chronotropic response and effective preload augmentation during exercise.^2,20^ In contrast, ccTGA is characterized by progressive atrioventricular conduction disease related to the abnormal position and course of the conduction system, resulting in a higher prevalence of complete heart block with advancing age.^1,4,21^ In our cohort, complete heart block was more common in ccTGA, whereas pacemaker implantation was more frequent among d-TGA patients, reflecting the distinct electrophysiologic substrates and pacing indications in these two anatomic groups. Despite these differences, both cohorts exhibited a high burden of electrical disease that plausibly contributes to reduced exercise tolerance.

Beyond structural and electrophysiologic considerations, our findings underscore the importance of psychological factors in shaping functional limitation among adults with sRV. In the combined cohort, patients with documented anxiety or depression demonstrated lower exercise capacity, worse NYHA functional class, and lower KCCQ scores. In contrast, BMI did not differ according to anxiety or depression status and was not associated with exercise capacity, suggesting that the observed relationship between mental health burden and functional limitation is unlikely to be explained by differences in body habitus alone. Notably, the association between mental health status and exercise capacity was most pronounced in d-TGA patients, whereas ccTGA patients showed less differentiation by psychological diagnosis. This pattern suggests that the alignment between perceived symptoms and objective physiologic limitation may be more apparent in atrial switch patients, potentially reflecting higher arrhythmia burden, greater chronotropic limitation, increased comorbidities, and/or cumulative exposure to medical interventions over the life course. Importantly, the attenuation of this relationship in multivariable analysis suggests that mental health may act as a correlated or modifying factor rather than an independent determinant of exercise capacity. Anxiety and depression may therefore amplify or reflect underlying physiologic limitation rather than directly driving it. The relationship between mental health burden and sRV systolic function further illustrates the complexity of this interaction. While anxiety was associated with lower sRV ejection fraction in the combined cohort, this relationship did not persist within anatomic subgroups, indicating that mental health status is unlikely to be a direct determinant of ventricular systolic performance.

Rather, psychological distress may modify functional capacity through behavioral and perceptual pathways, including reduced physical activity, diminished engagement in exercise or rehabilitation, heightened symptom awareness, and altered autonomic regulation. It is also possible that awareness of impaired sRV function and its associated complications contribute to increased anxiety. These mechanisms may interact bidirectionally with cardiac physiology, reinforcing a cycle in which physiologic impairment and psychological distress exacerbate one another over time.

The observed prevalence of anxiety and depression in our cohort appears higher than that reported in prior multicenter studies, including the APPROACH-IS II cohort, which demonstrated impaired quality of life among patients with a sRV compared with those with systemic left ventricles, with a substantial proportion of this effect mediated by ventricular dysfunction.^22^ Several factors may account for this difference. APPROACH-IS II was an international, cross-sectional study with heterogeneous patient populations and standardized survey-based assessments, whereas our single-center cohort reflects longitudinal clinical documentation within a tertiary referral setting, where patients may have more advanced disease and greater cumulative healthcare exposure. This finding may reflect the increasing mental health burden that accompanies aging with a sRV and prolonged exposure to medical and surgical interventions. In addition, clinically documented diagnoses may capture patients with more severe or persistent psychological distress, potentially enriching for a higher-risk subgroup. These differences highlight the importance of cohort context and ascertainment methods when interpreting the burden of mental health disease in adults with CHD.

From a clinical standpoint, these findings support a more integrated approach to the care of adults with a sRV. While ventricular function, arrhythmia surveillance, and heart failure therapies remain central to management, our data suggest that routine assessment of mental health and patient-reported outcomes provides complementary insight into functional limitation. Brief screening tools for anxiety and depression can be feasibly incorporated into ACHD clinic workflows and may help identify patients whose symptom burden and exercise intolerance are disproportionate to traditional imaging metrics.^23,24^ Addressing psychological distress alongside physiologic impairment may therefore represent an opportunity to improve both perceived and measured health status in this population.

More broadly, these results argue against viewing sRV dysfunction as a purely mechanical or electrophysiologic condition. Instead, adults with sRV appear to exhibit a coupled cardio-psychological phenotype in which anatomy, surgical history, ventricular adaptation, arrhythmia burden, and mental health collectively shape functional capacity and quality of life. At the same time, the dominant independent effect of anatomic subtype highlights the importance of underlying physiologic substrate in determining exercise performance, even after accounting for conventional clinical and imaging variables. Recognizing this integrated phenotype has important implications for both clinical care and future research, as interventions focused solely on ventricular mechanics may be insufficient to meaningfully improve outcomes without parallel attention to psychological well-being.

## Conclusions

Adults with sRV physiology exhibit a closely integrated physiologic and psychological phenotype. In this cohort, patients with d-TGA following atrial switch demonstrated lower exercise capacity and greater overall disease burden compared with those with ccTGA. Anatomic subtype emerged as the primary independent correlate of exercise capacity, whereas ventricular function and mental health were not independently associated after adjustment. These findings highlight the importance of considering the underlying physiologic substrate alongside patient-reported and psychological factors and support a more comprehensive approach to assessment that extends beyond traditional imaging metrics in adults with CHD.

Future studies should define and quantify the bidirectional relationship between declining cardiac function and rising mental health burden in adults with a sRV, with the goal of developing integrated, mind–body therapeutic strategies that target both physiologic impairment and psychological well-being. In addition, evaluation of other CPET parameters beyond ppVO2 may provide further mechanistic insight.

### Study Limitations

This study is limited by its retrospective design and single-center population. Mental health diagnoses were derived from clinical documentation rather than standardized psychological assessment and may therefore underestimate the true prevalence of anxiety and depression. Lower KCCQ quartiles contained small sample sizes, precluding robust statistical comparisons. Although clinical data were collected longitudinally, the analyses were cross-sectional and therefore cannot establish causality or temporal relationships. CMR data were incomplete for some participants. The multivariable analysis of KCCQ was additionally limited by missing patient-reported data, reducing statistical power to detect independent associations. The cohort size precluded robust sex-stratified analyses, limiting assessment of potential sex-based differences in the observed associations.

## Clinical Perspectives

### Clinical Implications

These findings support a comprehensive approach to evaluating adults with a sRV that extends beyond assessment of ventricular function alone. Lower exercise capacity was associated with anxiety, depression, worse functional status, and poorer health-related quality of life, while anatomic subtype remained the primary independent correlate of exercise capacity. These results support integrating cardiopulmonary exercise testing, advanced cardiac imaging, patient-reported health status, and routine mental health screening to improve phenotyping and guide multidisciplinary management of adults with sRV physiology.

### Translational Outlook

Prospective longitudinal studies are needed to determine whether changes in exercise capacity and mental health predict progression to heart failure, arrhythmias, hospitalization, transplantation, or death in adults with a sRV. Future investigations should evaluate whether multidisciplinary interventions targeting both physiologic limitation and psychological well-being improve functional capacity, quality of life, and long-term clinical outcomes.

## Data Availability

All data produced in the present work are contained in the manuscript

## Acknowledgements

The authors thank the Adult Congenital Heart Association for its support of this work through a research grant.

## Abbreviations

ACHD: adult congenital heart disease
ccTGA: congenitally corrected transposition of the great arteries
CHD: congenital heart disease
CMR: cardiac magnetic resonance
CPET: cardiopulmonary exercise testing
d-TGA: d-transposition of the great arteries
KCCQ: Kansas City Cardiomyopathy Questionnaire
NYHA: New York Heart Association
ppVO2: percent-predicted peak oxygen consumption
sRV: systemic right ventricle

